# ASSESSING THE ACCESSIBILITY, AFFORDABILITY, AND ACCEPTABILITY OF REFRACTIVE SERVICES AS BARRIERS TO UPTAKE OF THESE SERVICES IN KAKAMEGA MUNICIPALITY, KENYA

**DOI:** 10.1101/2024.07.24.24310925

**Authors:** Kisenge Masinde Martin, Ebrahim Khan Naimah

**Affiliations:** Lecturer Makerere University, Uganda; Lecturer University of KwaZulu Natal, South Africa

**Keywords:** Barriers, Accessibility, Affordability, Availability, Uptake, Refractive services

## Abstract

**Background information:** Refractive errors and presbyopia remain a burden to the entire population. An estimated 76% of the 191 million blind people have preventable or treatable causes. Uncorrected Refractive Error (URE), the number one cause (51%) of moderate and severe vision impairment is easily preventable.

**Aim:** The study aimed to evaluate the accessibility, affordability, and acceptability of refractive services in Kakamega Municipality.

**Methodology:** A population-based descriptive cross-sectional study was undertaken in Kakamega municipality using a cluster sampling method and descriptive data analysis.

**Results:** Out of 358 participants, 199 (55.6%) were male and 159 (44.4%) were female. The analysis shows affordability (18.3%) as the main reason for not using spectacles, followed by lack of quality care (3.4%), access to eye care (3.4%), awareness (2.5%), unpleasant past experiences (2.2%), importance not given to eye care issues (1.6%), lack of communication (0.9%), and disapproval from family members (0.9%). The study found that the affordable price range for spectacles varies between Kshs.5000 and less than Kshs.2000. More participants (38.0%) reported above Kshs.5000, while 29% indicated less Kshs.2000. The study found that affordability (p = 0.000), availability (p=0.004), and accessibility (p=0.005) of refractive services significantly influenced the uptake of these services.

**Conclusion:** The study reveals that refractive services in Kakamega municipality are not easily accessible due to the lack of adequate services in government hospitals. Additionally, patients in the municipality struggle to afford spectacles due to the direct cost of spectacles and the lack of services in easily accessible public facilities.

## Introduction

### Background and rationale

Uncorrected refractive errors are a significant cause of vision impairment and blindness, with the global average being 43% ^1^. In 2010, refractive errors were the second leading cause of blindness after cataracts^2^. The challenge lies in correcting these errors globally, particularly in Africa, where vision impairment is higher in developing countries like Kenya^1^. India and China account for approximately 50% of global vision impairment and blindness due to uncorrected refractive errors ^3, 4^. The annual global economic burden attributed to distance vision impairment due to uncorrected refractive errors is estimated at $220 billion, while the cost for training manpower and establishing service delivery facilities is only $28 billion ^5,6^.

Accessibility and affordability are key factors affecting the uptake of refractive services. Universal eye health is needed to provide 100% universal access to healthcare, which can be achieved by increasing coverage of services ^7^. Addressing uncorrected refractive errors requires human resource development, service delivery, social enterprise, infrastructure, and supplies ^8^. In Africa, there is an unequal provision of refractive training, which poses a challenge to maintaining uniformity in service quality. Integrating refraction services into existing healthcare systems is also necessary ^6, 9^. In Kenya, there is a limited number of eye care workers and inadequate human resource capacity in government institutions, leading patients to seek care at government institutions due to financial reasons ^10^.

## Research methods

This study used a population-based descriptive cross-sectional design to investigate refractive errors and presbyopia in a population aged 18-60 years in Kakamega town. The study used cluster sampling to select households in four administrative sub-locations, with subjects aged 18-60 years with vision below 6/12 that improved with pinhole testing included. The sample size was 384 people, selected based on Krejcie and Morgan’s table^11^. A questionnaire was used to interview participants with refractive errors and presbyopia identified through visual acuity testing. Data was analyzed using SPSS version 26, with frequencies and chi-square computed. The research was approved by the Kenyan ethical clearance committee and presented to the local government administration. The study followed procedures to ensure ethical clearance and data collection.

## Results

### Spectacle coverage among participants in Kakamega municipality

The study found that 55.4% of participants used spectacles for near reading, while 34.5% used them for far distance vision correction. Other reasons included general near vision (2.4%), light sensitivity (2.4%), and cosmetic reasons (0.4%). One hundred and nine participants did not provide reasons (Shown in table 1 below)

**Table 1.** Participants stating reasons for spectacle use.

| *Responses* | *f* | *Rel. f* | *cf* | *Percentile* |
| --- | --- | --- | --- | --- |
| *Cosmetic* | 1 | 0.003 | 358 | 100.00 |
| *for near vision* | 6 | 0.017 | 357 | 99.72 |
| *for near reading* | 138 | 0.385 | 351 | 98.04 |
| *for far distance* | 86 | 0.240 | 213 | 59.50 |
| *Others* | 8 | 0.022 | 127 | 35.47 |
| *don't know* | 4 | 0.011 | 119 | 33.24 |
| *light sensitivity* | 6 | 0.017 | 115 | 32.12 |
| *Did not respond to this question* | 109 | 0.304 | 109 | 30.45 |

### Perspectives of affordability, accessibility and availability of refractive services in Kakamega municipality

#### Reasons for not using spectacles

The analysis showed that a majority had no specific reason for not using spectacles(56.7%),affordability (18.3%) as the main reason for not using spectacles, followed by lack of quality care(3.4%), access to eye care (3.4%), awareness (2.5%), unpleasant past experiences (2.2%), importance not given to eye care issues (1.6%), lack of communication (0.9%), and disapproval from family members (0.9%) shown in table 2.

**Table 2.** Participants stating reasons for not using spectacles.

| ***Responses*** | ***f*** | ***Rel. f*** | ***cf*** | ***Percentile*** |
| --- | --- | --- | --- | --- |
| ***No reason*** | 203 | 0.567 | 358 | 100.00 |
| ***Affordability*** | 59 | 0.165 | 155 | 43.30 |
| ***Lack of quality care*** | 11 | 0.031 | 96 | 26.82 |
| ***Lack of access to eye care services*** | 11 | 0.031 | 85 | 23.74 |
| ***Lack of communication with others regarding health issues*** | 3 | 0.008 | 74 | 20.67 |
| ***Family disapproval or pressure*** | 3 | 0.008 | 71 | 19.83 |
| ***Importance not given to eye care issues*** | 5 | 0.014 | 68 | 18.99 |
| ***Unpleasant experience*** | 7 | 0.020 | 63 | 17.60 |
| ***Shyness*** | 2 | 0.006 | 56 | 15.64 |
| ***Don't know*** | 7 | 0.020 | 54 | 15.08 |
| ***Others*** | 1 | 0.003 | 47 | 13.13 |
| ***Lack of awareness*** | 8 | 0.022 | 46 | 12.85 |
| ***Problem was corrected*** | 1 | 0.003 | 38 | 10.61 |
| ***Refusal to use*** | 1 | 0.003 | 37 | 10.34 |
| ***Did not respond to this question*** | 36 | 0.101 | 36 | 10.06 |

#### Affordable price range for spectacles in Kakamega Municipality

The study found that the affordable price range for spectacles varies between Kshs.5000 and less than Kshs.2000. More participants (38.0%) reported above Kshs.5000, while 29% indicated less Kshs.2000. This difference may be due to the main types of spectacles worn: near vision and distance vision. Reading spectacles are cheaper than distance vision correction spectacles. The types of spectacles were not investigated (figure 1).

**Figure 1.**
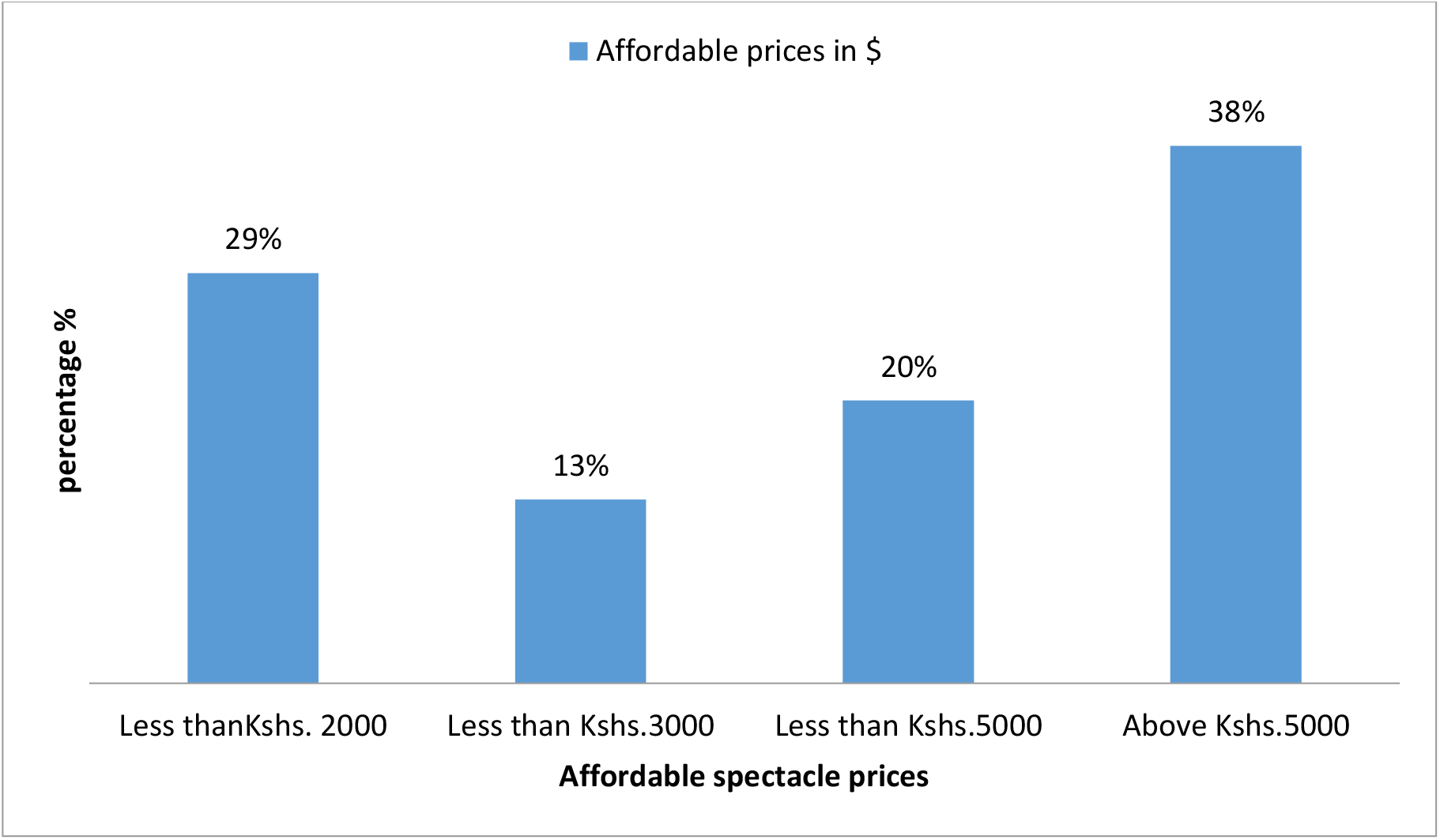
Participant perceptions regarding an affordable price for spectacles.

#### Factors for not using/discontinuation of wearing spectacles in children

The study revealed that 42.9% of children discontinue spectacle use due to breakage and carelessness, followed by the cost (35.4%). Other reasons include discomfort (4.2%), personal beliefs (9.1%), lack of access to eye facilities (2.3%), fear of use (1.6%), ignorance (1.3%), peer pressure (1.0%), and a 0.3% report that their vision improved, indicating that spectacles may no longer be necessary (figure 2 below).

**Figure 2.**
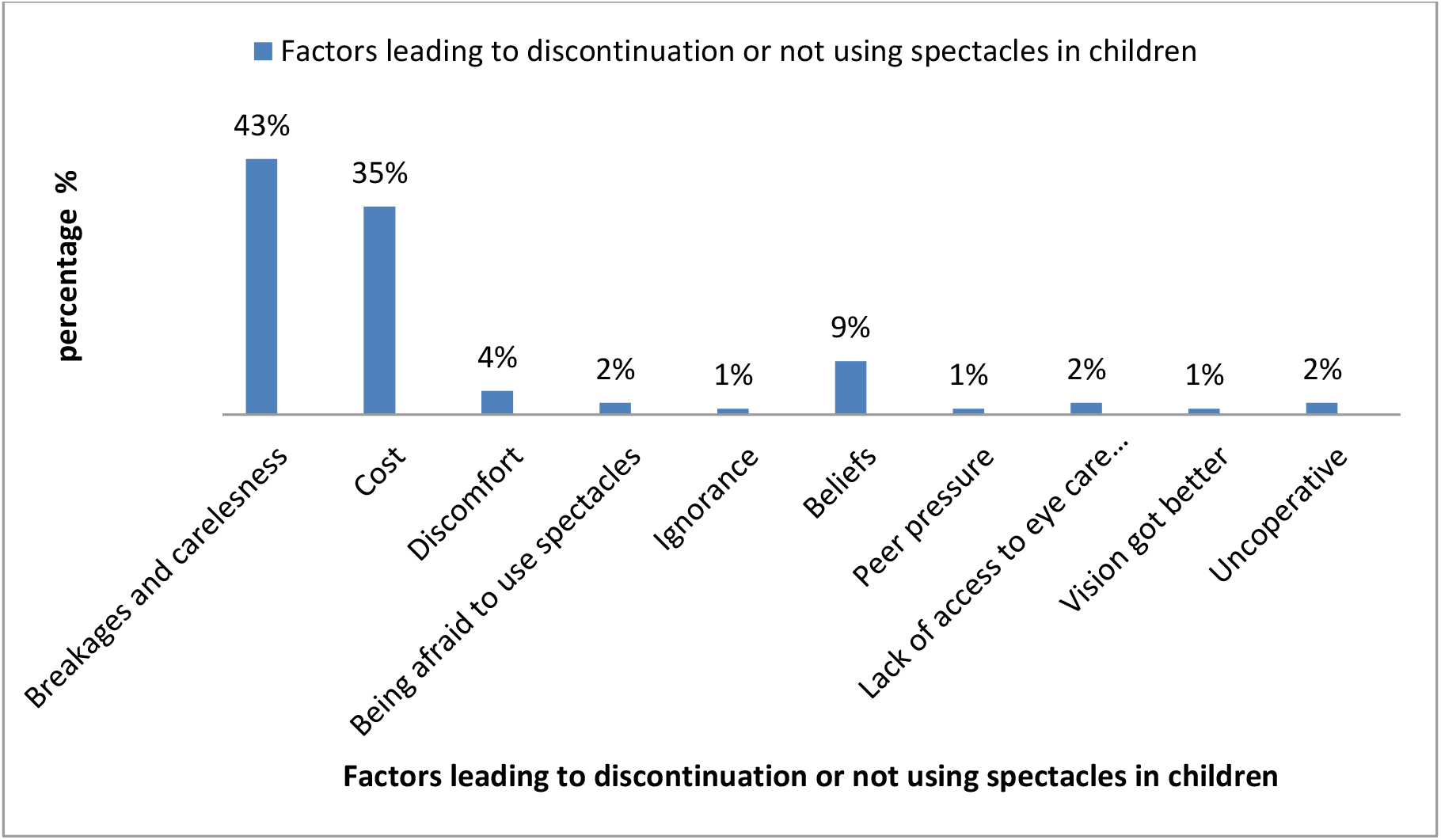
Reasons for not using/discontinuation of wearing spectacles in children.

### A Chi-square analysis was conducted to assess the affordability, availability, and accessibility of refractive services in Kakamega municipality

The study found that affordability, availability, and accessibility of refractive services significantly influenced the uptake of these services, rejecting the null hypothesis (shown in table 3 below)

**Table 3.** Chi-square analysis was done on affordability, availability and accessibility of refractive services in Kakamega municipality.

| ***Variables*** | **Coefficient value** | **P-value** |
| --- | --- | --- |
| ***Affordability of refractive services*** | 0.233 | 0.000 |
| ***Availability of refractive services*** | 0.173 | 0.004 |
| ***Accessibility of refractive services*** | 0.189 | 0.005 |

## Discussions

The low uptake of refractive services in Kenya is attributed to affordability and perception towards spectacle-wearing. Near reading glasses are cheap and readily available, leading many participants to use them. Presbyopia is the most highly corrected refractive error, and addressing barriers to refractive errors will result in good coverage of spectacles. Affordable prices for spectacles and integrated knowledge about eye care within the health system and through community engagement could be solutions to these barriers^12^.

The majority of the sample population seeks health services from government hospitals, with private hospitals/clinics being second. Government hospitals or health centers in Kenya do not include eye units as part of their services, and those with eye units are not well equipped to provide optical services. This hinders the uptake of refractive services in the Kakamega Municipality. Many members of the community resort to seeking eye care at private optical shops, which are often more expensive and sometimes unaffordable ^13^.

A developed economy approach to deliver refractive and eye services, such as those in Europe, South Africa, Australia, New Zealand, and North America, can greatly improve the uptake of refractive services within the country. The affordability of refractive services is significantly associated (p=0.000) with the use of spectacles, with the cheapest spectacles being Kshs.5000 ^12^. To tackle this challenge, the government should equip public hospitals to provide optical services at a lower rate, and bulk purchase of frames and lenses at reduced prices will help decrease prices and reduce the burden of avoidable blindness and poor vision^12^. Additionally, availability and accessibility of refractive services are significantly associated (p=0.004 and p=0.005 respectively) with low uptake, as most participants receive eye services from government hospitals but do not offer optical services, making accessibility and availability a barrier to the same services.

## Conclusions

The study reveals that refractive services in Kakamega municipality are not easily accessible due to the lack of adequate services in government hospitals. Additionally, patients in the municipality struggle to afford spectacles due to the direct cost of spectacles and the lack of services in easily accessible public facilities.

### Study limitations

The study aimed to interview 371 participants, but only 358 were interviewed due to locked houses and family members away from the home, possibly due to the study being conducted during working days and most people were at their workplaces.

### Recommendations

The ministry of public health can use the findings of this research to advocate for the integration of refractive services into community health facilities, ensuring that they are adequately staffed and equipped to meet the needs of patients. They should also consider including refractive services in the national health insurance fund for all citizens, regardless of employment status. Bulk purchases of consumables like spectacle frames and lenses can help reduce costs and address affordability barriers. Health care providers should provide accurate information about refractive services to patients addressing misconceptions. The research should be published in public health journals and other sources to serve as a reference for scholars interested in similar studies in different settings.

## Data Availability

All data produced in the present work are contained in the manuscript

## List of abbreviations

URE: Uncorrected refractive errors.
VI: Visual impairment
WHO: World health organization.
RE: Refractive err

## Definition of terms

A **Refractive error:** occurs when the image is not focused on the retina ^14^.

A **Barrier:** is something or reason that makes someone not use or access something ^12^.

**Visual impairment:** is when someone has unaided visual acuity less than 6/12 in a better eye ^12^.

**Uncorrected refractive error:** is when a person has visual acuity of less than 6/12 but improves to 6/12 or more on the use of a pinhole ^12, 3^.

**Presbyopia:** it is when a person has a near vision of less than N8 with both eyes open at a normal working distance in an individual that is above 35 years^3^

